# Autosomal Dominant Polycystic Kidney Disease does not significantly alter major COVID-19 outcomes among veterans

**DOI:** 10.1101/2020.11.25.20238675

**Authors:** Xiangqin Cui, Julia W. Gallini, Christine L. Jasien, Michal Mrug

## Abstract

Chronic kidney disease (CKD), as well as its common causes (e.g., diabetes and obesity), are recognized risk factors for severe COVID-19 illness. To explore whether the most common inherited cause of CKD, autosomal dominant polycystic kidney disease (ADPKD), is also an independent risk factor, we studied data from the VA health system and the VA COVID-19-shared resources (e.g., ICD codes, demographics, pre-existing conditions, pre-testing symptoms, and post-testing outcomes). Among 61 COVID-19-positive ADPKD patients, 21 (34.4%) were hospitalized, 10 (16.4%) were admitted to ICU, 4 (6.6%) required ventilator, and 4 (6.6%) died by August 18, 2020. These rates were comparable to patients with other cystic kidney diseases and cystic liver-only diseases. ADPKD was not a significant risk factor for any of the four outcomes in multivariable logistic regression analyses when compared with other cystic kidney diseases and cystic liver-only diseases. In contrast, diabetes was a significant risk factor for hospitalization [OR 2.30 (1.61, 3.30), p<0.001], ICU admission [OR 2.23 (1.47, 3.42), p<0.001], and ventilator requirement [OR 2.20 (1.27, 3.88), p=0.005]. Black race significantly increased the risk for ventilator requirement [OR 2.00 (1.18, 3.44), p=0.011] and mortality [OR 1.60 (1.02, 2.51), p=0.040]. We also examined the outcome of starting dialysis after COVID-19 confirmation. The main risk factor for starting dialysis was CKD [OR 6.37 (2.43, 16.7)] and Black race [OR 3.47 (1.48, 8.1)]. After controlling for CKD, ADPKD did not significantly increase the risk for newly starting dialysis comparing with other cystic kidney diseases and cystic liver-only diseases. In summary, ADPKD did not significantly alter major COVID-19 outcomes among veterans when compared to other cystic kidney and liver patients.

## Introduction

According to the US Center for Disease Control (CDC), people of any age with chronic kidney disease (CKD) are at increased risk of severe illness from COVID-19 ^1^. In support of this statement, a meta-analysis of four early studies from China concluded that CKD is a significant risk factor for severe COVID-19 symptoms ^2^, although individual studies failed to detect the significance of this relationship. Similarly, CKD was a significant risk factor for in-hospital mortality (OR 2.14) in the univariable analysis of 200 COVID-19 positive patients admitted to a tertiary New York medical center; however, this association was not significant in the multivariable analysis ^3^. CKD was also a significant risk factor for pneumonia (OR 2.05) and mortality (OR 2.48) among 23,162 COVID-19 positive patients from Mexico after adjusting for age, gender, and smoking status ^4^. In addition, CKD was an independent significant risk factor for hospital visits but not for respiratory support requirements among the people who tested positive for COVID-19, according to an analysis of over 2 million COVID-19 smart app users ^5^. Finally, the estimated hazard ratio (HR) of CKD as a risk factor for COVID-19-related death was 1.3 for eGFR at 30-60 and 2.5 for eGFR<30 after controlling for all other major comorbidities in a recent study of over 17 million participants in the UK’s NHS in England ^6^.

In the majority of CKD patients, the etiology of CKD is complex. The underlying medical condition to which CKD is attributed often leads to multiorgan manifestations, which may further contribute to the development and progression of CKD. For example, obesity is a strong risk factor for the onset of CKD and its progression. Further, obesity increases the risk for developing diabetes mellitus type 2 and hypertension, the two leading causes of CKD (reviewed by Kovesdy et al. ^7^). Obesity is also an independent risk factor for the activity of autoimmune disorders, the third leading cause of CKD. Notably, obesity, diabetes mellitus type 2, and immunosuppression (commonly used for the treatment of autoimmune disorders) are recognized by CDC together with CKD among the eight conditions that increase the risk of severe illness from COVID-19 at any age ^1^. The CDC also recognizes hypertension as a factor that may increase the risk of severe illness from COVID-19 ^1^. Such a complex relationship among major risk factors for CKD and COVID-19 outcomes complicates the assessment of direct effects of CKD on COVID-19 complications. While existing studies provide strong support for CKD and its causes as key risk factors for major COVID-19 complications, they do not clearly delineate between the risk attributable to CKD itself and the risk attributable to its underlying causes.

However, some CKD causes affect mainly the kidneys and have less impact on other organ systems compared to other causes of CKD. Such more kidney-specific causes include a subset of inherited disorders. Among them is CKD’s most common inherited form, autosomal dominant polycystic kidney disease (ADPKD; MIM: 173900 and 613095). ADPKD is the fourth leading cause of CKD and end-stage kidney disease in the US. With the prevalence of 4.3:10,000, ADPKD affects over 140,000 people in the US ^8^ and over 13 million people with ADPKD worldwide ^9^. Progressively worsening cystic renal disease and renal function loss are the hallmark ADPKD manifestations, though some people with ADPKD may also develop extrarenal complications such as liver cysts, vascular (e.g., intracranial) aneurysms, and heart valve abnormalities. Therefore, ADPKD is a leading cause of CKD that allows for assessment of CKD’s direct effects on major COVID-19 outcomes with limited confounding by other medical complications.

To explore the impact of ADPKD on COVID-19 outcomes, we studied participants in the largest integrated US health system, the Veterans Health Administration (VHA), which serves more than 9 million veterans each year. Specifically, we analyzed the effects of ADPKD on major COVID-19 outcomes, including hospitalization, ICU admission, ventilator requirement, and mortality. We compared these outcomes in ADPKD COVID-19 positive and negative veterans with other (non-ADPKD) cystic renal disease patients (e.g., simple renal cysts) and patients with cystic liver disease (without renal cysts).

## Materials and Methods

### Study cohort

We created a VA cystic kidney and liver cohort using electronic health record data from the VA Corporate Data Warehouse (CDW). We restricted this cohort to patients diagnosed between 1/1/2000 and 1/31/2020. After that, we identified CKD patients by extracting relevant clinical and demographic data for all patients with ICD10 and ICD9 codes for cystic kidney disease (Q61 and 753.1, respectively). A subset of these patients was identified as ADPKD-enriched by using the ICD10 code for Polycystic kidney, adult type (Q61.2) and ICD9 codes for Polycystic kidney disease - autosomal dominant and Polycystic kidney (disease) unspecified (753.13 and 753.12). We designated the remaining (non-ADPKD cystic kidney) patients as the Other cystic kidney group. We then selected patients with ICD10 and ICD9 codes for the cystic disease of the liver (Q44.6 and 751.62, respectively) to serve as controls. Patients with multiple ICD codes were included in only one disease group with priority given to the ADPKD-enriched group followed by the Other cystic kidney group. Patients who had only ICD codes for the cystic disease of the liver were assigned to the Cystic liver-only group. In summary, we analyzed three groups derived from the VA cystic kidney and liver cohort: the ADPKD-enriched group, and its two controls, the Other cystic kidney group, and the Cystic liver-only group.

This study was approved by the Emory University Institutional Review Board (IRB00115069) and Atlanta VA medical center R&D committee (2019-111232).

### COVID-19 status

The VA Informatics and Computing Infrastructure (VINCI) maintains and updates VA COVID-19 shared resources, including data for all the VA patients and employees that were tested or suspected for COVID-19 (https://www.hsrd.research.va.gov/covid19.cfm). VINCI also provides patient data on demographic characteristics, pre-existing conditions, pre-testing symptoms, post-testing conditions, and post-testing procedures. The COVID-19 shared data resources are updated weekly. After obtaining required administrative approvals, including from the VA Institutional Review Board, we received access to the COVID-19 shared resource for patients from the above described VA Cystic kidney and liver cohort. We analyzed the data available from VINCI as of August 18, 2020.

### Studied variables

Data on four main COVID-19 outcomes (hospitalization, ICU admission, ventilator requirement, and mortality) were ascertained 60 days after each patient’s COVID-19 test. All four outcomes were measured as yes/no. Given the ongoing data collection, about half of the patients had not yet reached 60 days after COVID-19 testing. However, since the outcomes analyzed are most likely to occur shortly after testing, the underestimation of the outcome rate should be low. The definitions of analyzed variables by the VINCI COVID-19 shared resource are listed in **Supplementary Table 1**.

Demographic data were obtained mainly from the VA Observational Medical Outcomes Partnership (OMOP) datasets, a consolidated and well-curated data source ^10^. The pre-existing conditions for potential COVID-19 risk factors were extracted from the VA electronic medical records for the two years prior to COVID-19 testing unless specified otherwise. BMI was calculated from the available height and weight data closest to the testing date.

### Statistical Analysis

Summary statistics were generated using the R package *TableOne*. P values were from Fisher’s exact test or χ^2^ test for categorical variables (depending on sample size) and the Wilcoxon rank sum test for continuous variables. The multivariable logistic regression was conducted using only patients with non-missing outcomes. Race was consolidated into two categories, Black/African American and Other. We performed a sensitivity analysis where we treated missing outcome values as negative outcomes for the multivariable logistic regression. To better handle censoring, we also conducted multivariable cox regressions using time to the outcome as the dependent variable instead of measuring each outcome as yes/no. Since timing data among COVID-19 negative patients was limited, our multivariable Cox regression was only conducted for the COVID-19 positive patients. R packages *finalfit, ggplot2* and *jtools* were used for results summary and visualization.

## Results

### ADPKD-enriched cohort

In the ADPKD-enriched group derived from the VA cystic kidney and liver cohort, 709 veterans were tested for COVID-19; 61 of them were positive. In comparison, 9817 veterans from the Other cystic kidney disease group were tested for COVID-19; 803 were positive. In the Cystic liver-only group, 788 veterans were tested for COVID-19; 52 were positive (**Figure 1**).

**Figure 1.**
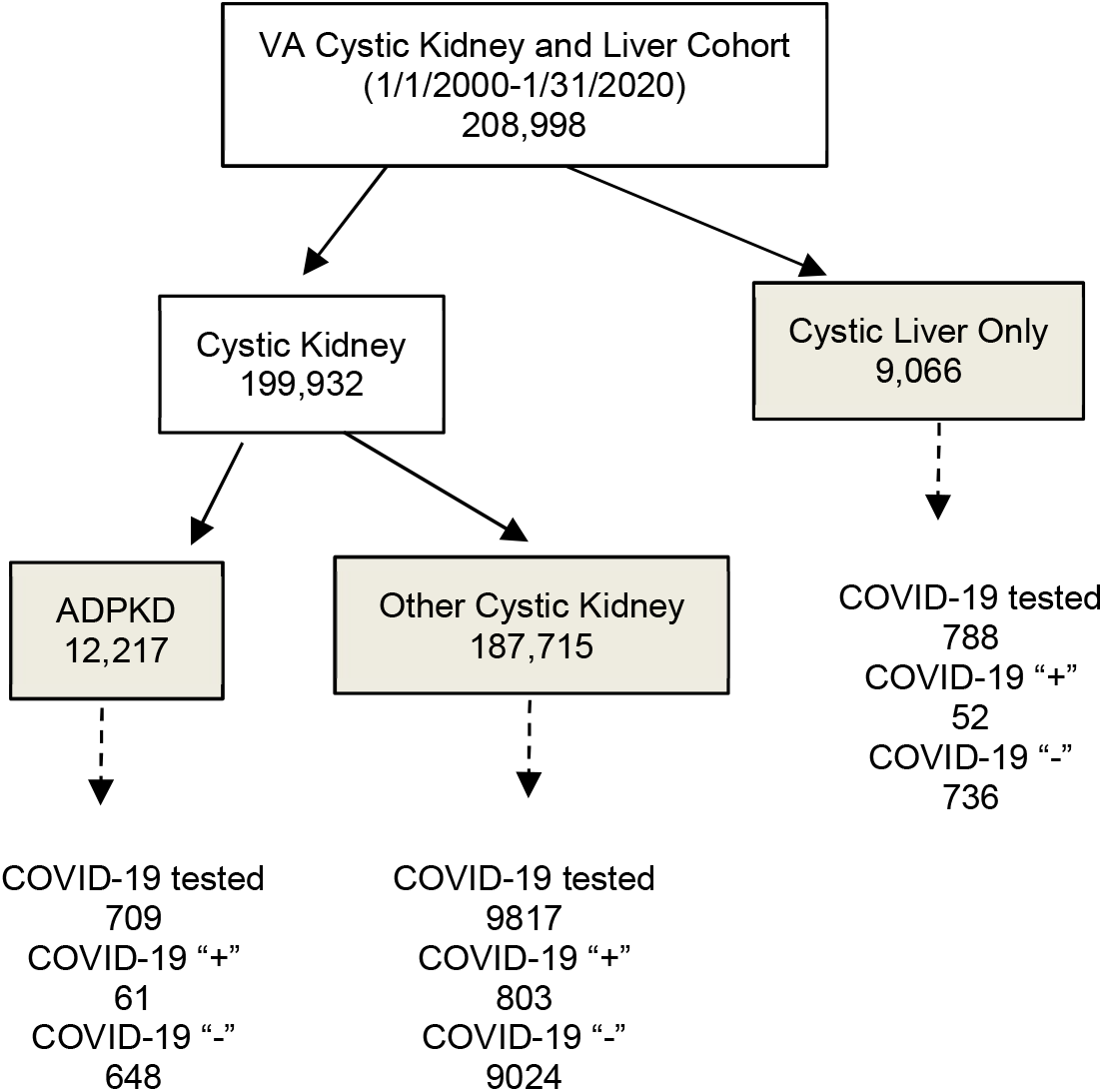
VA Cystic kidney and liver study cohort groups extracted from VA electronic medical records. COVID-19 test results were obtained from VA COVID-19 Shared Resource. “+”, positive; “-”, negative

### Demographic characteristics and pre-existing conditions

The distribution of demographic characteristics of COVID-19-positive patients in the studied cohort closely resembles the VA patient population, which is dominated by older males (**Table 1**). The median age of patients in the ADPKD-enriched subset of this cohort was 5-6 years lower than the median age of the Other cystic kidney group subset and the Cystic liver-only group subset (66, 72, and 71 years, respectively). As expected, the patients in the ADPKD-enriched group had a higher rate of kidney-related pre-existing conditions (e.g., acute kidney failure, chronic kidney disease, and dialysis requirement) than those from the Other cystic kidney and the Cystic liver-only groups.

**Table 1.** Demographic characteristics and pre-existing conditions of the COVID-19 positive patients

| **Variables** | **Other cystic kidney** | **Cystic liver-only** | **ADPKD** | **p** |
| --- | --- | --- | --- | --- |
| **n** | 803 | 52 | 61 |  |
| **Sex = Male (%)** | 768 (95.6) | 46 (88.5) | 58 (95.1) | 0.064 |
| **Race (%)** |  |  |  | 0.802 |
| American Indian or Alaska Native | 3 ( 0.4) | 0 ( 0.0) | 1 ( 1.6) |  |
| Asian | 3 ( 0.4) | 0 ( 0.0) | 0 ( 0.0) |  |
| Black or African American | 305 (38.0) | 23 (44.2) | 24 (39.3) |  |
| Native Hawaiian or Other Pacific Islander | 6 ( 0.7) | 0 ( 0.0) | 0 ( 0.0) |  |
| Unknown | 38 ( 4.7) | 1 ( 1.9) | 1 ( 1.6) |  |
| White | 448 (55.8) | 28 (53.8) | 35 (57.4) |  |
| **Age (median [IQR])** | 72.0 [65.0, 78.0] | 71.0 [61.8, 76.0] | 65.0 [54.0, 74.0] | <0.001 |
| **BMI (median [IQR])** | 28.7 [25.4, 33.0] | 30.3 [25.4, 33.5] | 28.1 [25.3, 32.2] | 0.712 |
| **Chronic Lung Disease (%)** | 322 (40.1) | 24 (46.2) | 22 (36.1) | 0.548 |
| **Cancer (%)** | 288 (35.9) | 17 (32.7) | 18 (29.5) | 0.559 |
| **Any Diabetes (%)** | 386 (48.1) | 15 (28.8) | 24 (39.3) | 0.014 |
| **Liver Disease (%)** | 86 (10.7) | 18 (34.6) | 5 ( 8.2) | <0.001 |
| **Active Liver Injury (%)** | 3 ( 0.4) | 0 ( 0.0) | 0 ( 0.0) | 0.809 |
| **AKF (%)** | 137 (17.1) | 2 ( 3.8) | 16 (26.2) | 0.006 |
| **CKF (%)** | 63 ( 7.8) | 1 ( 1.9) | 19 (31.1) | <0.001 |
| **CKD (%)** | 301 (37.5) | 12 (23.1) | 45 (73.8) | <0.001 |
| **Dialysis (%)** | 61 ( 7.6) | 1 ( 1.9) | 19 (31.1) | <0.001 |
| **Dialysis At Testing (%)** | 22 ( 2.7) | 1 ( 1.9) | 8 (13.1) | <0.001 |

There is no missing data for any of these variables. IQR, inter quartile range; AKF, acute kidney failure; CKF, chronic kidney failure; CKD, chronic kidney disease.

We also noted that among the COVID-19-positive patients, there was a relatively higher proportion of African Americans (10% higher) and a lower rate of chronic lung disease within each disease group when compared to COVID-19-negative patients in this study (**Supplementary Table 2**).

### Pre-testing COVID-19 symptoms

We compared COVID-19-associated symptoms in the 30 days prior to COVID-19 testing among patients who tested positive (**Table 2**). All of the studied COVID-19 symptoms were comparable across the ADPKD-enriched, Other cystic kidney disease, and Cystic liver-only groups. ADPKD patients had a lower risk of cough; however, this association did not reach statistical significance. Among all the COVID-19 positive patients studied, about 48% were asymptomatic, and 30-40% had a fever. No patients lost their sense of smell or taste.

**Table 2.** COVID-19 symptoms within 30 days before the COVID-19 testing

| **Variables** | **Cystic Kidney** | **Cystic Liver** | **ADPKD** | **p** |
| --- | --- | --- | --- | --- |
| n | 803 | 52 | 61 |  |
| Abdominal Pain (%) | 24 ( 3.0) | 2 ( 3.8) | 3 ( 4.9) | 0.68 |
| Chills (%) | 16 ( 2.0) | 0 ( 0.0) | 0 ( 0.0) | 0.318 |
| Cold (%) | 79 ( 9.8) | 7 ( 13.5) | 5 ( 8.2) | 0.626 |
| Cough (%) | 146 ( 18.2) | 8 ( 15.4) | 4 ( 6.6) | 0.064 |
| Diarrhea (%) | 49 ( 6.1) | 5 ( 9.6) | 3 ( 4.9) | 0.542 |
| Dyspnea (%) | 124 ( 15.4) | 5 ( 9.6) | 12 ( 19.7) | 0.334 |
| Fatigue (%) | 75 ( 9.3) | 4 ( 7.7) | 3 ( 4.9) | 0.48 |
| Fever (%) | 256 ( 31.9) | 12 ( 23.1) | 23 ( 37.7) | 0.246 |
| Headache (%) | 18 ( 2.2) | 0 ( 0.0) | 1 ( 1.6) | 0.53 |
| Loss Of Smell (%) | 0 ( 0.0) | 0 ( 0.0) | 0 ( 0.0) | --- |
| Loss Of Taste (%) | 0 ( 0.0) | 0 ( 0.0) | 0 ( 0.0) | --- |
| Myalgia (%) | 10 ( 1.2) | 1 ( 1.9) | 1 ( 1.6) | 0.892 |
| Nausea (%) | 21 ( 2.6) | 2 ( 3.8) | 4 ( 6.6) | 0.198 |
| No Symptoms (%) | 386 ( 48.1) | 25 ( 48.1) | 29 ( 47.5) | 0.997 |
| Rhinorrhea (%) | 1 ( 0.1) | 0 ( 0.0) | 0 ( 0.0) | 0.932 |
| Sore Throat (%) | 8 ( 1.0) | 1 ( 1.9) | 0 ( 0.0) | 0.583 |

There is no missing data for these variables

### Major COVID-19 related outcomes

We evaluated four COVID-19 outcomes in the 60 days after testing: hospitalization, ICU admission, Ventilator requirement, and mortality (**Figure 2** and **Supplementary Table 3**).

**Figure 2.**
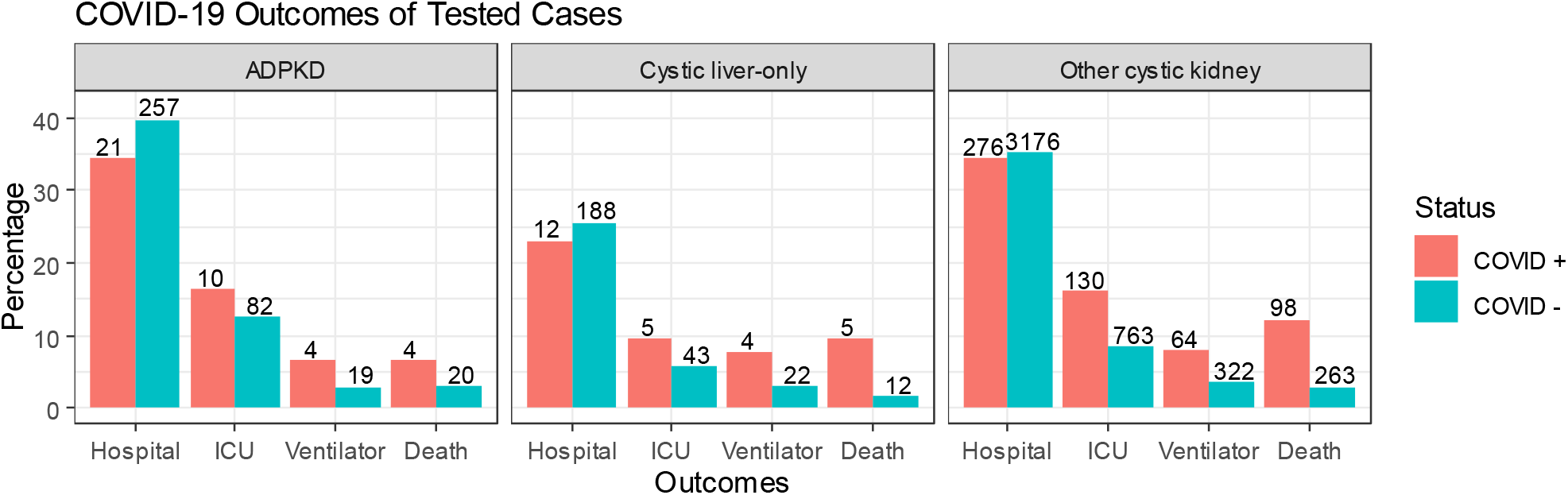
Percentage of patients with major COVID-19 related outcomes among patients tested positive and negative separately. The number of patients is labeled on top of each bar.

The hospitalization rate was similar between COVID-19 positive and negative patients for both ADPKD-enriched and Other cystic kidney disease groups (34-40%); it was about 25% for both COVID-19 positive and negative patients in the Cystic liver-only group. The lack of an increase in hospitalization rate among the COVID-19 positive (vs. negative) patients across all three groups was likely related to hospitalized patients’ priority testing during the study period.

The ICU admission rate in the 60 days after testing was higher among the COVID-19 positive (vs. negative) patients across all three disease groups. About 16% of COVID-19 positive patients in the ADPKD-enriched and the Other cystic kidney disease group were admitted to the ICU compared to 10% in the Cystic liver-only group. COVID-19 positive Other cystic kidney patients had a significantly higher rate of ICU admission than their COVID-19 negative counterparts. The respective rates in COVID-19 positive vs. negative patients were not significantly different among patients in the ADPKD-enriched or Cystic liver-only groups, likely due to the sample size (**Supplementary Table 3**).

The rate of ventilator requirement among COVID-19 positive patients (8%-9%) was double of that among COVID-19 negative patients (3%-4%); the significance was reached only in the Other cystic kidney disease group (not the ADPKD-enriched and Cystic liver-only groups due to smaller sample size). The incidence rates were similar among the three groups derived from the studied VA cohort.

Death rate of COVID-19 positive patients was significantly higher when compared to COVID-19 negative patients (death rate 10% vs 2-3%).

Similar results were obtained when the above analyses were done on hospitalized patients alone (**Supplementary Figure 1**). We performed such analyses to make our data more comparable to earlier studies that analyzed only hospitalized patients.

### Multivariable logistic regression failed to establish ADPKD as an independent risk factor

We used two multivariable logistic regression models (one for COVID-19 positive patients and one for negative patients) to analyze the four outcomes of interest. Disease status (ADPKD vs. Other cystic kidney vs. Cystic liver-only) was the main exposure. Models were adjusted for pre-existing conditions, especially known risk factors for severe COVID-19 symptoms. Specifically, we controlled for age, body mass index (BMI), prior CKD, diabetes, dialysis, cancer, and liver problems (see the odds ratio (OR) and 95% confidence interval for all predictor variables in **Figure 3**). ADPKD did not achieve statistical significance as an independent risk factor for any of the four COVID-19 outcomes (hospitalization, ICU admission, ventilator requirement, and mortality). However, an OR of greater than one was estimated for hospitalization (1.37), ICU admission (1.72), and ventilator requirement (1.51).

**Figure 3.**
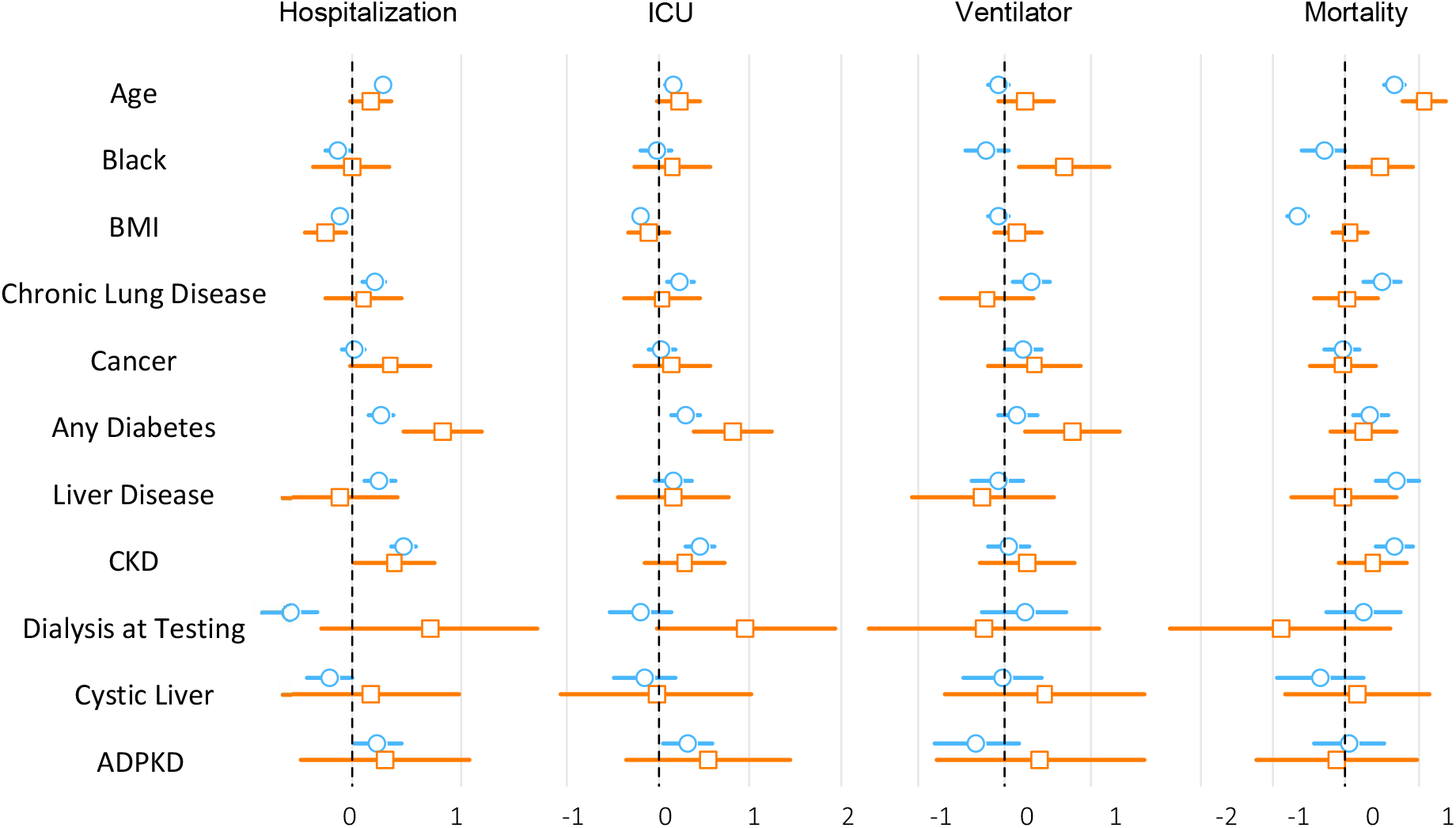
Multivariable logistic regression odds ratio and 95% confidence intervals reported on a logarithmic scale for COVID-19 positive (orange) and negative (blue) patients. Outcomes include hospitalization, ICU admission, ventilator requirement, and mortality. All tested patients were considered, and missing outcomes were removed.

### Effects of comorbidities on COVID-19 outcomes

Some of the known COVID-19 risk factors showed significant effects in our analysis. Diabetes was the most prominent independent risk factor with an OR of 2.30 (1.61, 3.30) for hospitalization, 2.23 (1.47, 3.42) for ICU admission, and 2.20 (1.27, 3.88) for ventilator requirement. Other significant risk factors include being Black/African American for ventilator requirement with OR 2.0 (1.18, 3.44) and age for death with OR 1.10 (1.07, 1.12) where the unit for age was one year.

### Effects of COVID-19 on new start of dialysis

Since kidney injury has been demonstrated to be a consequence of COVID-19 infection ^11,12^, we evaluated the dialysis status change after COVID-19 testing (**Supplementary Table 3)**. No change was found in the majority of patients. Very few patients stopped dialysis (≤ 0.6%). A small percentage of patients started dialysis after their COVID-19 test (7-8% in ADPKD patients, 2-4% in cystic kidney patients, and <1% in cystic liver disease). We conducted a multivariable logistic regression to evaluate the risk of starting dialysis among patients who were not on dialysis at the time of their COVID-19 test (**Figure 4** and **Table 3)**. Due to the small number of new dialysis cases, we combined the cystic liver and cystic kidney as the reference group for ADPKD. The most significant risk factor for new dialysis was pre-existing chronic kidney disease (CKD), OR 6.37 (2.43-16.7) for COVID-19 positive patients. The relationship is even more prominent for COVID-19 negative patients [OR 72.99 (26.83, 198.57)], which could be related to much lower risk in the non-CKD patients that were COVID-19 negative. In addition, Black race was also a risk factor with OR 3.47 (1.48, 8.1) for COVID-19 positive patients and OR 1.82 (1.35, 2.46) for COVID-19 negative patients. ADPKD was a significant risk factor among COVID-19 negative patients, OR 2.36 (1.62, 3.45) and among COVID-19 positive patients, OR 3.48 (0.97, 12.4) did not reach statistical significance.

**Table 3.**
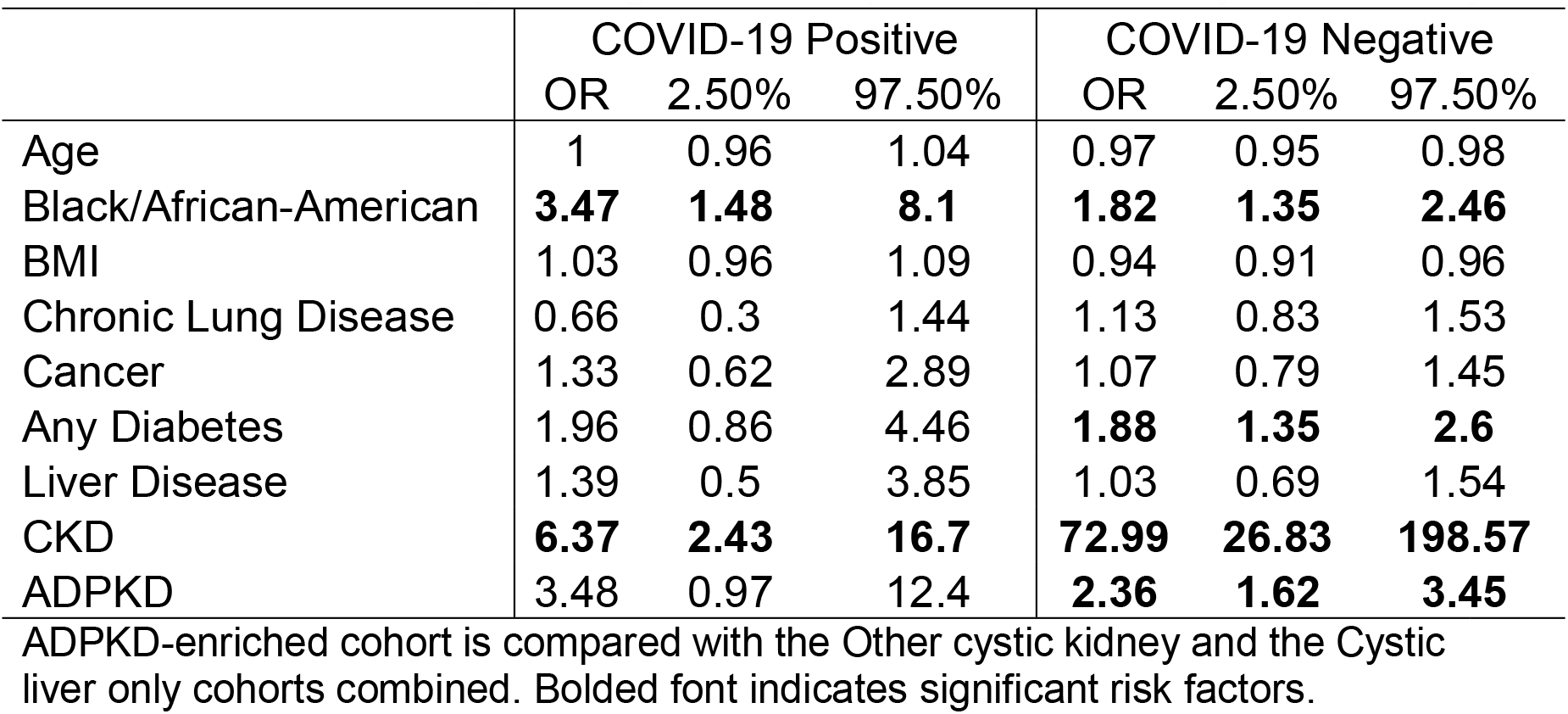
Multivariable logistic regression for patients who newly started dialysis

**Figure 4.**
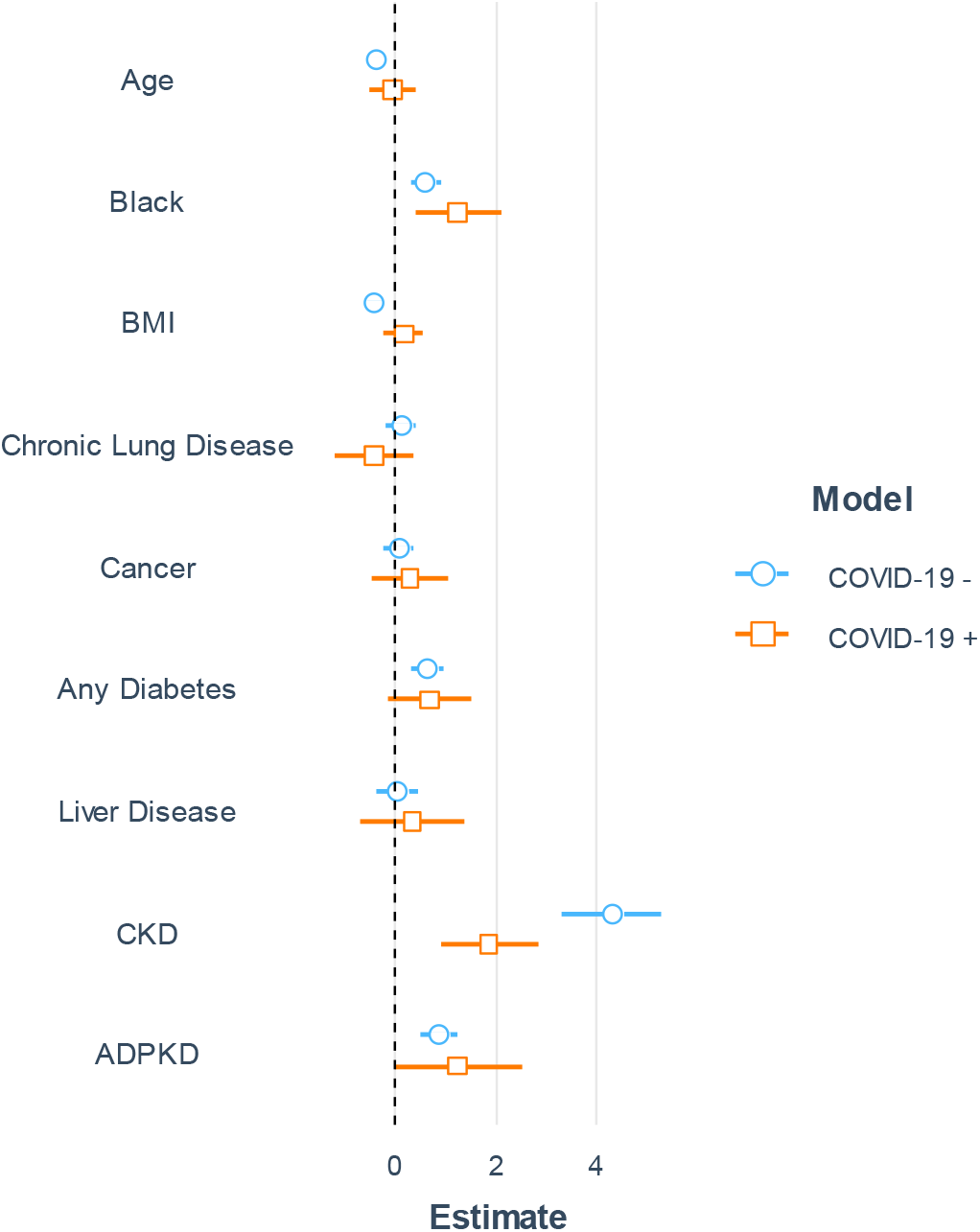
Multivariable logistic regression analysis for newly dialysis gained after COVID-19 testing. Only the patients not on dialysis at testing were used for analysis. The odds ratio is reported on a logarithmic scale.

## Discussion

The large patient population in the VA health system allowed evaluation of ADPKD as an independent risk factor for severe illness from COVID-19 infection. Among the 61 COVID-19 positive ADPKD patients, the rates of hospitalization, ICU admission, ventilator requirement, and mortality were comparable to those from patients with Other cystic kidney disease and Cystic liver-only disease. These rates were consistently higher than those from the COVID-19 negative patients as expected; although, statistical significance was not reached in most comparisons due to the small sample size. The ICU admission rate among hospitalized ADPKD patients that were COVID-19 positive was similar to the 48.5% seen among male COVID-19 positive patients in a Kaiser Permanente study of 1840 hospital admitted COVID-19 positive patients ^13^. The death rate among hospitalized COVID-19 positive patients is also comparable (around 20% in both studies). The lack of added adverse effects of ADPKD on COVID-19 complications demonstrated in this study suggests that ADPKD does not significantly exacerbate a patient’s risk for severe COVID-19 illness compared to patients with other cystic kidney diseases or liver diseases.

The validity of the data and analyses described in this manuscript is supported by identifying established risk factors for severe COVID-19 illness as statistically significant in the studied VA cystic kidney and liver cohort. This includes identifying diabetes as an independent risk factor for hospitalization, ICU admission, and ventilator use. Our analyses also identified Black race and age as risk factors for ventilator requirement and mortality. In addition, consistent with CKD’s earlier recognition as the central risk factor for acute kidney injury ^14^, we identified CKD as the leading risk factor for new dialysis among COVID-19 positive patients in the studied VA cohort.

Though our study had an adequate sample size to reveal the effects of factors with a major impact on the severe COVID-19 outcomes (diabetes, Black race), this study is underpowered to detect smaller effects from ADPKD. Similarly, the relatively small number of confirmed COVID-19 positive patients from the ADPKD-enriched group did not allow for stratification into CKD stages and end-stage kidney disease management options (e.g., kidney transplant vs. hemodialysis vs. peritoneal dialysis). Therefore, we plan to repeat the reported analyses in the future when data for more COVID-19 ADPKD patients are available. These future analyses will allow confirmation of the reported associations and provide more granular data analyses, such as stratifying CKD based on underlying causes for risk comparison and mediation analyses to delineate the direct and indirect risk contributions.

Another limitation of the current study is that COVID-19 tested patients were not necessarily representative samples of the CKD and cystic liver population. Instead, the patients that were hospitalized or admitted to the ICU were preferably tested. This is a likely explanation for why the hospitalization rate and ICU rate in the studied VA cohort were relatively high. Thus, the rate of ventilator requirement and death rate likely better reflect the severity of COVID-19 infection in this cohort. In addition, we did not have a healthy control group in our cohort study. We only compared the patients among the three groups: ADPKD-enriched, Other cystic kidney disease, and Cystic liver-only disease. Our results for ADPKD are relative to the other two groups.

An additional limitation was the relatively large amount of missing data for outcomes because many patients had not yet reached 60 days since testing. The missing data were left out of some rate calculations, which likely resulted in an overestimation of the rates. Patients who have gone weeks (though short of 60 days) without being hospitalized, admitted to the ICU, etc., are unlikely to reach these outcomes by the 60-day mark, so while these data were considered missing, they were not missing at random.

Another limitation was the use of ICD codes to assign diagnoses in the studied VA cystic kidney and liver cohort. Although ICD codes are commonly used for disease phenotyping based on electronic medical records, their accuracy is disease-dependent and often inferior to machine learning-based complex methods ^15^. While it would be ideal to have diagnoses among these patients confirmed by genotyping, imaging, and family history data, such a depth of information will unlikely be fully retrievable from general medical records. Perhaps, future disease-centered registries might facilitate the correct assignment of diagnoses to patients in large datasets.

Finally, the data reported in this manuscript should be interpreted in the context of the VA population (∼75% White, 13% Black or African American, and 8% Hispanic or Latino) that consists predominantly of males (∼90%) ^16^.

In summary, we studied major outcomes (hospitalization, ICU admission, ventilator requirement, and death rates) of confirmed COVID-19 positive and COVID-19 negative patients among veterans who were assigned to groups classified as either ADPKD-enriched, Other cystic kidney disease, or Cystic liver-only disease. ADPKD, CKD type that mostly affects the kidneys, was not a significant independent risk factor for any of the four outcomes in multiple logistic regression analyses. Diabetes was a risk factor for hospitalization, ICU admission, and ventilator requirement; the Black race increased the risk for ventilator requirement and mortality. For both COVID-19 positive and negative patients, the leading risk factor for new dialysis treatment initiation was CKD and Black race. After controlling for CKD, ADPKD was not a significant independent risk factor for starting dialysis. Together, this study suggests that ADPKD is not a major risk for studied COVID-19 outcomes among veterans. However, this initial study will require validation when larger numbers of COVID-19 positive patients become available.

## Data Availability

The data are available in VA EHR databases. They are accessible to people with VA research credentials and approved for this study.

## Authors Contribution

X.C., J.G., C.J. and M.M. conceived and designed the research; X.C., J.G., C.J. and M.M. performed the experiments and carried out the analyses; X.C., J.G., C.J. and M.M. analyzed the data; X.C., J.G., C.J. and M.M. interpreted the results of experiments; X.C., C.J. prepared the figures and tables; X.C. and M.M. drafted the manuscript; X.C., J.G., C.J. and M.M. edited and revised the manuscript; X.C., J.G., C.J. and M.M. approved the final version of manuscript.

## Acknowledgements

Support was provided in part by the PKD Foundation research grant 247G20a (to X.C.), Atlanta VA Medical Center Research Office (X.C., J.W.G, C.J.), and by the National Institutes of Health (NIH)-funded University of Alabama at Birmingham (UAB) Hepato/Renal Fibrocystic Disease Core Center P30 DK074038 and Childhood Cystic Kidney Disease Core Center U54DK126087, the 1-I01-BX004232-01A2 grant from the Office of Research and Development, Medical Research Service, Department of Veterans Affairs and by the Detraz Endowed Research Fund in Polycystic Kidney Disease (to M. M.).

## Supplementary Material

**Supplementary Table 1.**
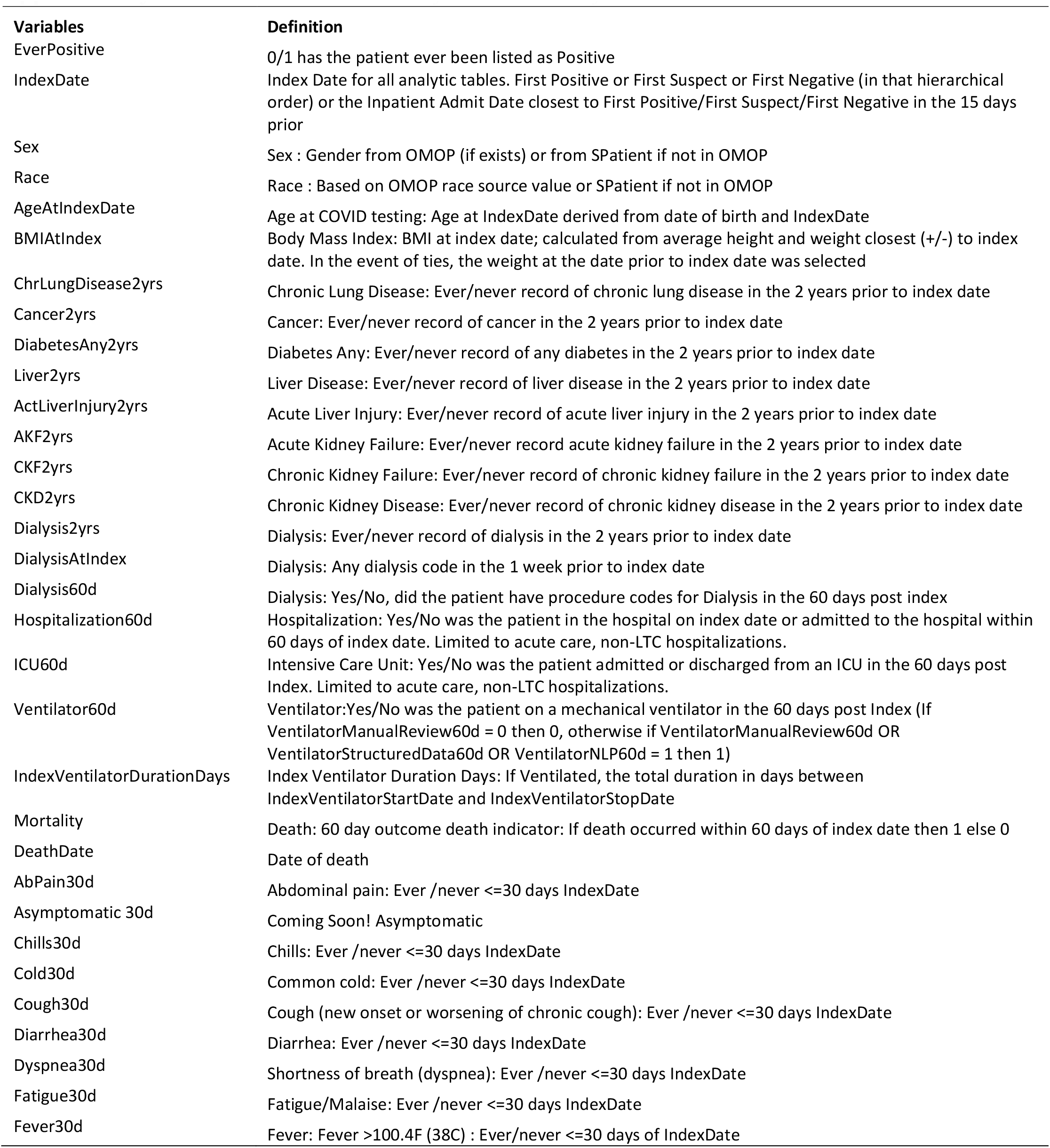
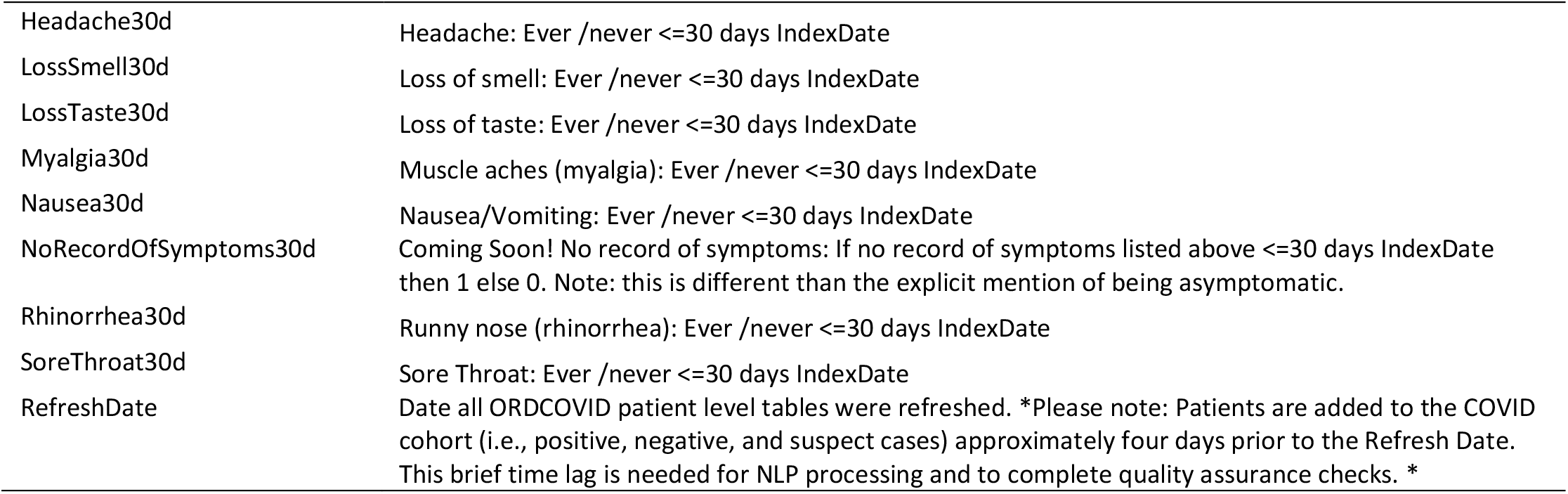
Definitions of analyzed variables by the VINCI COVID-19 shared resource.

| Variables | Definition |
| --- | --- |
| EverPositive | 0/1 has the patient ever been listed as Positive |
| IndexDate | Index Date for all analytic tables. First Positive or First Suspect or First Negative (in that hierarchical order) or the Inpatient Admit Date closest to First Positive/First Suspect/First Negative in the 15 days prior |
| Sex | Sex : Gender from OMOP (if exists) or from SPatient if not in OMOP |
| Race | Race : Based on OMOP race source value or SPatient if not in OMOP |
| AgeAtIndexDate | Age at COVID testing: Age at IndexDate derived from date of birth and IndexDate |
| BMIAtIndex | Body Mass Index: BMI at index date; calculated from average height and weight closest (+/-) to index date. In the event of ties, the weight at the date prior to index date was selected |
| ChrLungDisease2yrs | Chronic Lung Disease: Ever/never record of chronic lung disease in the 2 years prior to index date |
| Cancer2yrs | Cancer: Ever/never record of cancer in the 2 years prior to index date |
| DiabetesAny2yrs | Diabetes Any: Ever/never record of any diabetes in the 2 years prior to index date |
| Liver2yrs | Liver Disease: Ever/never record of liver disease in the 2 years prior to index date |
| ActLiverInjury2yrs | Acute Liver Injury: Ever/never record of acute liver injury in the 2 years prior to index date |
| AKF2yrs | Acute Kidney Failure: Ever/never record acute kidney failure in the 2 years prior to index date |
| CKF2yrs | Chronic Kidney Failure: Ever/never record of chronic kidney failure in the 2 years prior to index date |
| CKD2yrs | Chronic Kidney Disease: Ever/never record of chronic kidney disease in the 2 years prior to index date |
| Dialysis2yrs | Dialysis: Ever/never record of dialysis in the 2 years prior to index date |
| DialysisAtIndex | Dialysis: Any dialysis code in the 1 week prior to index date |
| Dialysis60d | Dialysis: Yes/No, did the patient have procedure codes for Dialysis in the 60 days post index |
| Hospitalization60d | Hospitalization: Yes/No was the patient in the hospital on index date or admitted to the hospital within 60 days of index date. Limited to acute care, non-LTC hospitalizations. |
| ICU60d | Intensive Care Unit: Yes/No was the patient admitted or discharged from an ICU in the 60 days post Index. Limited to acute care, non-LTC hospitalizations. |
| Ventilator60d | Ventilator: Yes/No was the patient on a mechanical ventilator in the 60 days post Index (If VentilatorManualReview60d = 0 then 0, otherwise if VentilatorManualReview60d OR VentilatorStructuredData60d OR VentilatorNLP60d = 1 then 1) |
| IndexVentilatorDurationDays | Index Ventilator Duration Days: If Ventilated, the total duration in days between IndexVentilatorStartDate and IndexVentilatorStopDate |
| Mortality | Death: 60 day outcome death indicator: If death occurred within 60 days of index date then 1 else 0 |
| DeathDate | Date of death |
| AbPain30d | Abdominal pain: Ever /never <=30 days IndexDate |
| Asymptomatic 30d | Coming Soon! Asymptomatic |
| Chills30d | Chills: Ever /never <=30 days IndexDate |
| Cold30d | Common cold: Ever /never <=30 days IndexDate |
| Cough30d | Cough (new onset or worsening of chronic cough): Ever /never <=30 days IndexDate |
| Diarrhea30d | Diarrhea: Ever /never <=30 days IndexDate |
| Dyspnea30d | Shortness of breath (dyspnea): Ever /never <=30 days IndexDate |
| Fatigue30d | Fatigue/Malaise: Ever /never <=30 days IndexDate |
| Fever30d | Fever: Fever >100.4F (38C) : Ever/never <=30 days of IndexDate |
| Headache30d | Headache: Ever /never <=30 days IndexDate |
| LossSmell30d | Loss of smell: Ever /never <=30 days IndexDate |
| LossTaste30d | Loss of taste: Ever /never <=30 days IndexDate |
| Myalgia30d | Muscle aches (myalgia): Ever /never <=30 days IndexDate |
| Nausea30d | Nausea/Vomiting: Ever /never <=30 days IndexDate |
| NoRecordOfSymptoms30d | Coming Soon! No record of symptoms: If no record of symptoms listed above <=30 days IndexDate then 1 else 0. Note: this is different than the explicit mention of being asymptomatic. |
| Rhinorrhea30d | Runny nose (rhinorrhea): Ever /never <=30 days IndexDate |
| SoreThroat30d | Sore Throat: Ever /never <=30 days IndexDate |
| RefreshDate | Date all ORDCOVID patient level tables were refreshed. *Please note: Patients are added to the COVID cohort (i.e., positive, negative, and suspect cases) approximately four days prior to the Refresh Date. This brief time lag is needed for NLP processing and to complete quality assurance checks. * |

**Supplementary Table 2.** Demographics and pre-existing conditions for patients in the VA study cohort that were tested COVID-10 negative

|  | Other cystic kidney | Cystic liver-only | ADPKD | p |
| --- | --- | --- | --- | --- |
| n | 8623 | 705 | 617 |  |
| Sex = M (%) | 8146 ( 94.5) | 614 ( 87.1) | 561 ( 90.9) | <0.001 |
| Race (%) |  |  |  | 0.001 |
| American Indian or Alaska Native | 67 ( 0.8) | 5 ( 0.7) | 2 ( 0.3) |  |
| Asian | 55 ( 0.6) | 10 ( 1.4) | 4 ( 0.6) |  |
| Black or African American | 2178 ( 25.3) | 205 ( 29.1) | 185 ( 30.0) |  |
| Native Hawaiian or Other Pacific Islander | 61 ( 0.7) | 11 ( 1.6) | 5 ( 0.8) |  |
| Unknown | 395 ( 4.6) | 40 ( 5.7) | 22 ( 3.6) |  |
| White | 5867 ( 68.0) | 434 ( 61.6) | 399 ( 64.7) |  |
| Age (median [IQR]) | 72.00 [65.00, 77.00] | 69.00 [61.00, 74.00] | 69.00 [59.00, 74.00] | <0.001 |
| BMI (median [IQR]) | 28.83 [25.11, 33.16] | 28.42 [24.86, 32.30] | 27.79 [24.17, 32.01] | <0.001 |
| ActLiverInjury = YES (%) | 36 ( 0.4) | 2 ( 0.3) | 1 ( 0.2) | 0.552 |
| AKF = YES (%) | 1619 ( 18.8) | 68 ( 9.6) | 168 ( 27.2) | <0.001 |
| Cancer = YES (%) | 3834 ( 44.5) | 313 ( 44.4) | 227 ( 36.8) | 0.001 |
| ChrLungDisease = YES (%) | 4293 ( 49.8) | 346 ( 49.1) | 277 ( 44.9) | 0.062 |
| LiverDisease = YES (%) | 1193 ( 13.8) | 212 ( 30.1) | 81 ( 13.1) | <0.001 |
| CKF = YES (%) | 684 ( 7.9) | 13 ( 1.8) | 221 ( 35.8) | <0.001 |
| CKD = YES (%) | 3230 ( 37.5) | 130 ( 18.4) | 453 ( 73.4) | <0.001 |
| DiabetesAny = YES (%) | 4202 ( 48.7) | 198 ( 28.1) | 235 ( 38.1) | <0.001 |
| Dialysis = YES (%) | 622 ( 7.2) | 12 ( 1.7) | 212 ( 34.4) | <0.001 |
| DialysisAtIndex = YES (%) | 265 ( 3.1) | 4 ( 0.6) | 77 ( 12.5) | <0.001 |

Note: The sample size of the COVID-19 negative patients is more than 10 times larger than that of positive patients.

**Supplementary Table 3.** Major outcomes for COVID-19 positive and negative patients.

| COVID-19 | ADPKD |  |  | Other cystic kidney |  |  | Cystic liver-only |  |  |
| --- | --- | --- | --- | --- | --- | --- | --- | --- | --- |
|  | negative | positive | p | negative | positive | p | negative | positive | p |
| n | 648 | 61 |  | 9014 | 803 |  | 736 | 52 |  |
| Dialysis status Change (%) |  |  | 0.816 |  |  | 0.001 |  |  | 1 |
| stopped | 4 ( 0.6) | 0 ( 0.0) |  | 26 ( 0.3) | 3 ( 0.4) |  | 0 ( 0.0) | 0 ( 0.0) |  |
| no change | 353 (54.5) | 23 (37.7) |  | 4596 (51.0) | 426 (53.1) |  | 343 (46.6) | 26 (50.0) |  |
| new | 49 ( 7.6) | 5 ( 8.2) |  | 154 ( 1.7) | 29 ( 3.6) |  | 3 ( 0.4) | 0 ( 0.0) |  |
| NA | 242 (37.3) | 33 (54.1) |  | 4238 (47.0) | 345 (43.0) |  | 390 (53.0) | 26 (50.0) |  |
| Hospitalization (%) |  |  | 0.507 |  |  | 0.651 |  |  | 0.818 |
| no | 183 (28.2) | 14 (23.0) |  | 2718 (30.2) | 269 (33.5) |  | 236 (32.1) | 16 (30.8) |  |
| yes | 257 (39.7) | 21 (34.4) |  | 3176 (35.2) | 276 (34.4) |  | 188 (25.5) | 12 (23.1) |  |
| NA | 208 (32.1) | 26 (42.6) |  | 3120 (34.6) | 258 (32.1) |  | 312 (42.4) | 24 (46.2) |  |
| ICU (%) |  |  | 0.528 |  |  | <0.001 |  |  | 0.424 |
| no | 315 (48.6) | 16 (26.2) |  | 4156 (46.1) | 356 (44.3) |  | 318 (43.2) | 20 (38.5) |  |
| yes | 82 (12.7) | 10 (16.4) |  | 763 ( 8.5) | 130 (16.2) |  | 43 ( 5.8) | 5 ( 9.6) |  |
| NA | 251 (38.7) | 35 (57.4) |  | 4095 (45.4) | 317 (39.5) |  | 375 (51.0) | 27 (51.9) |  |
| Ventilator (%) |  |  | 0.25 |  |  | <0.001 |  |  | 0.152 |
| no | 363 (56.0) | 19 (31.1) |  | 4465 (49.5) | 396 (49.3) |  | 330 (44.8) | 21 (40.4) |  |
| yes | 19 ( 2.9) | 4 ( 6.6) |  | 322 ( 3.6) | 64 ( 8.0) |  | 22 ( 3.0) | 4 ( 7.7) |  |
| NA | 266 (41.0) | 38 (62.3) |  | 4227 (46.9) | 343 (42.7) |  | 384 (52.2) | 27 (51.9) |  |
| Ventilator Duration Days (median [IQR]) | 2.50 [2.00, 4.25] | 11.00 [8.00, 12.00] | 0.079 | 3.00 [2.00, 7.00] | 10.00 [5.00, 18.50] | <0.001 | 3.00 [3.00, 4.00] | 3.00 [3.00, 8.50] | 0.634 |
| Mortality (%) | 20 ( 3.1) | 4 ( 6.6) | 0.288 | 263 ( 2.9) | 98 (12.2) | <0.001 | 12 ( 1.6) | 5 ( 9.6) | 0.001 |

Note: The p values are for testing the positive rates of each outcome, where missing data were treated as negative. Dialysis status change is from the comparison of the pre-testing to 60 days post testing. NA, missing.

**Supplementary Table 4.**
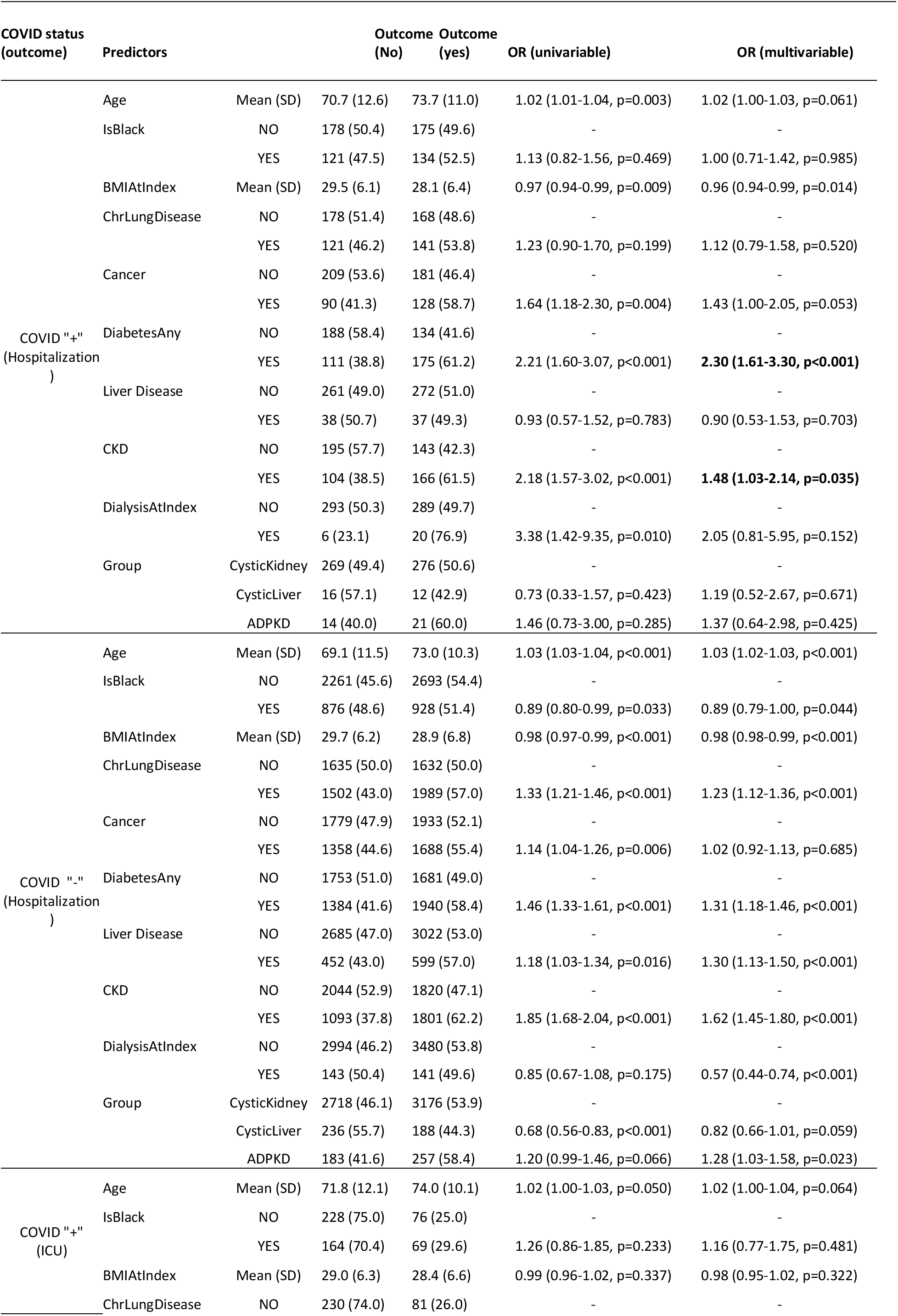
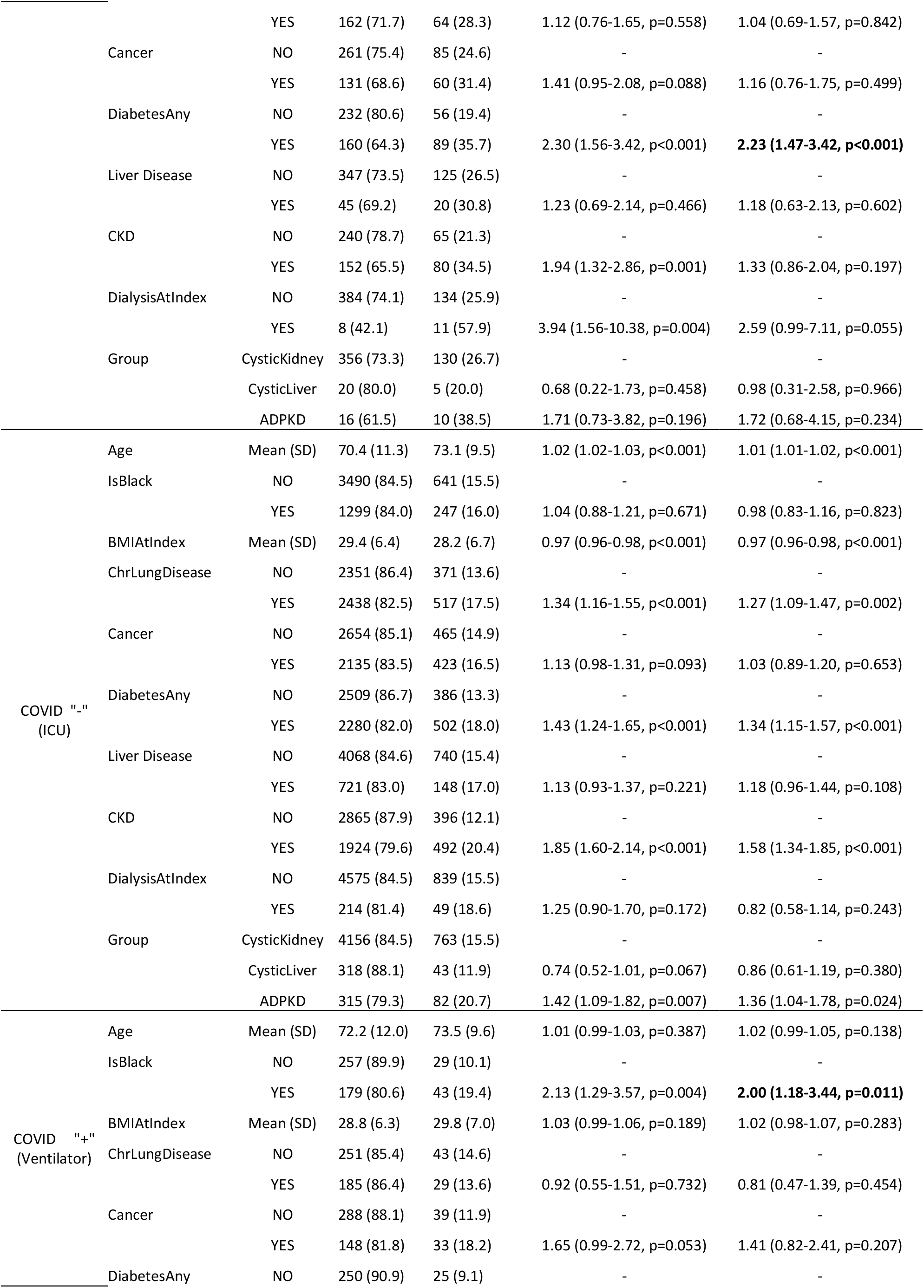
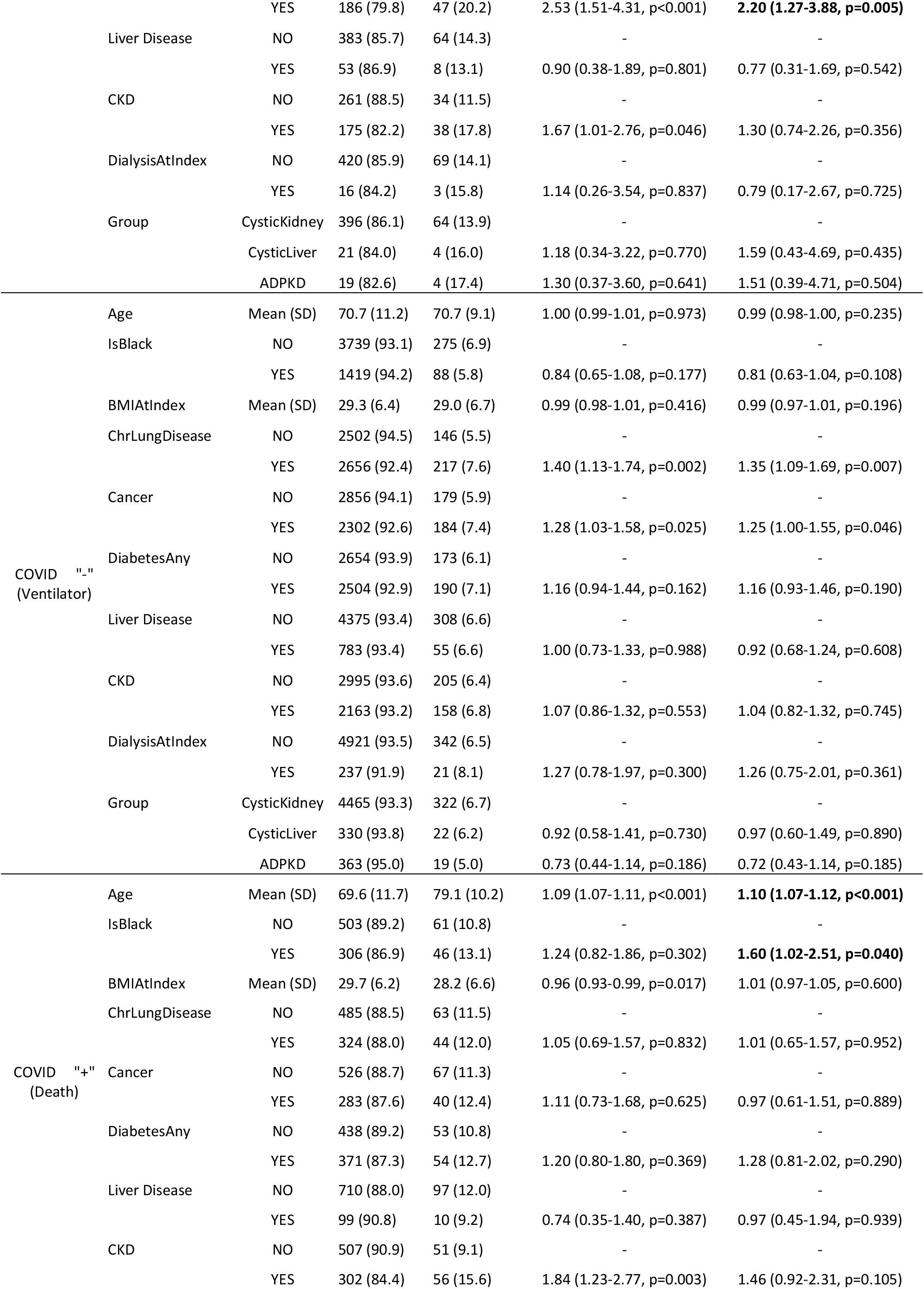
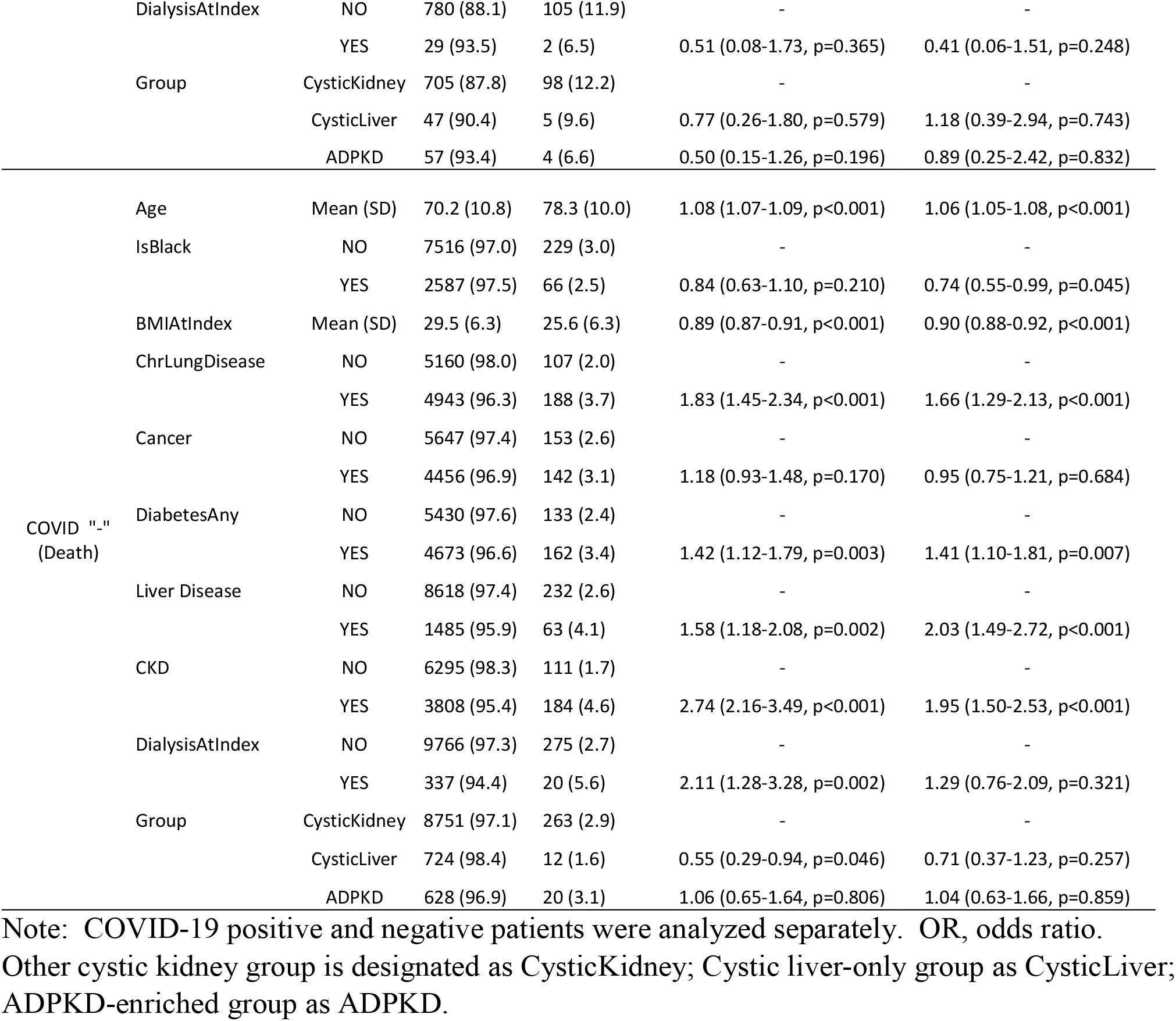
Univariable and multivariable logistic regression results

**Supplementary Table 5.**
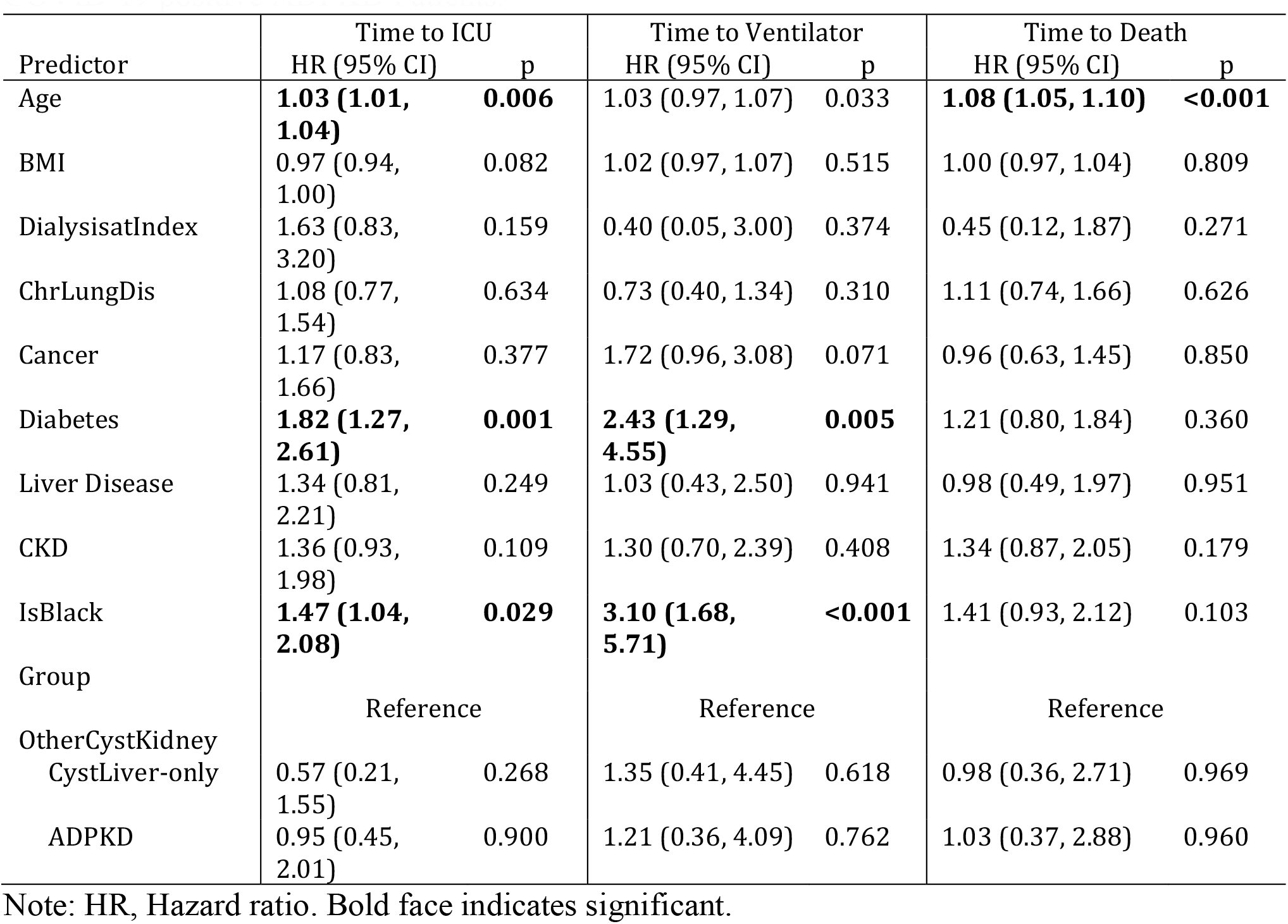
Time to Event analysis results using Cox regression for Outcomes in COVID-19 positive ADPKD Patients.

**Supplementary Figure 1.**
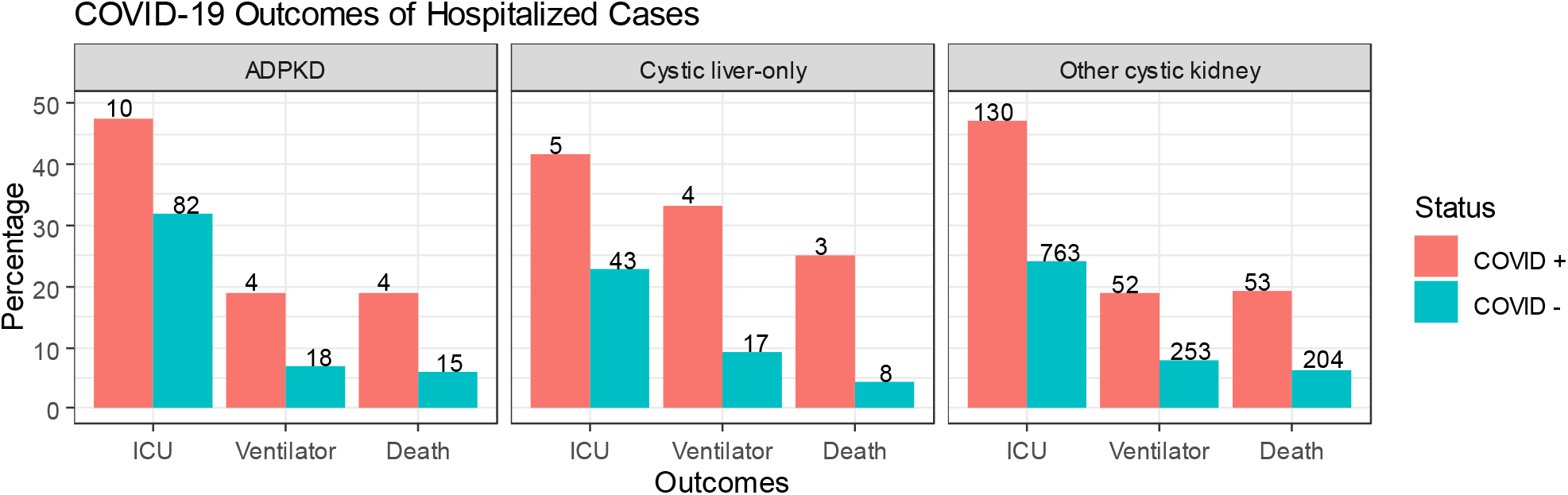
Percentage of patients with major COVID-19 outcomes in positive cases and negative cases among hospitalized patients. The number of patients is provided at the top of each bar.

**Supplementary Figure 2.**
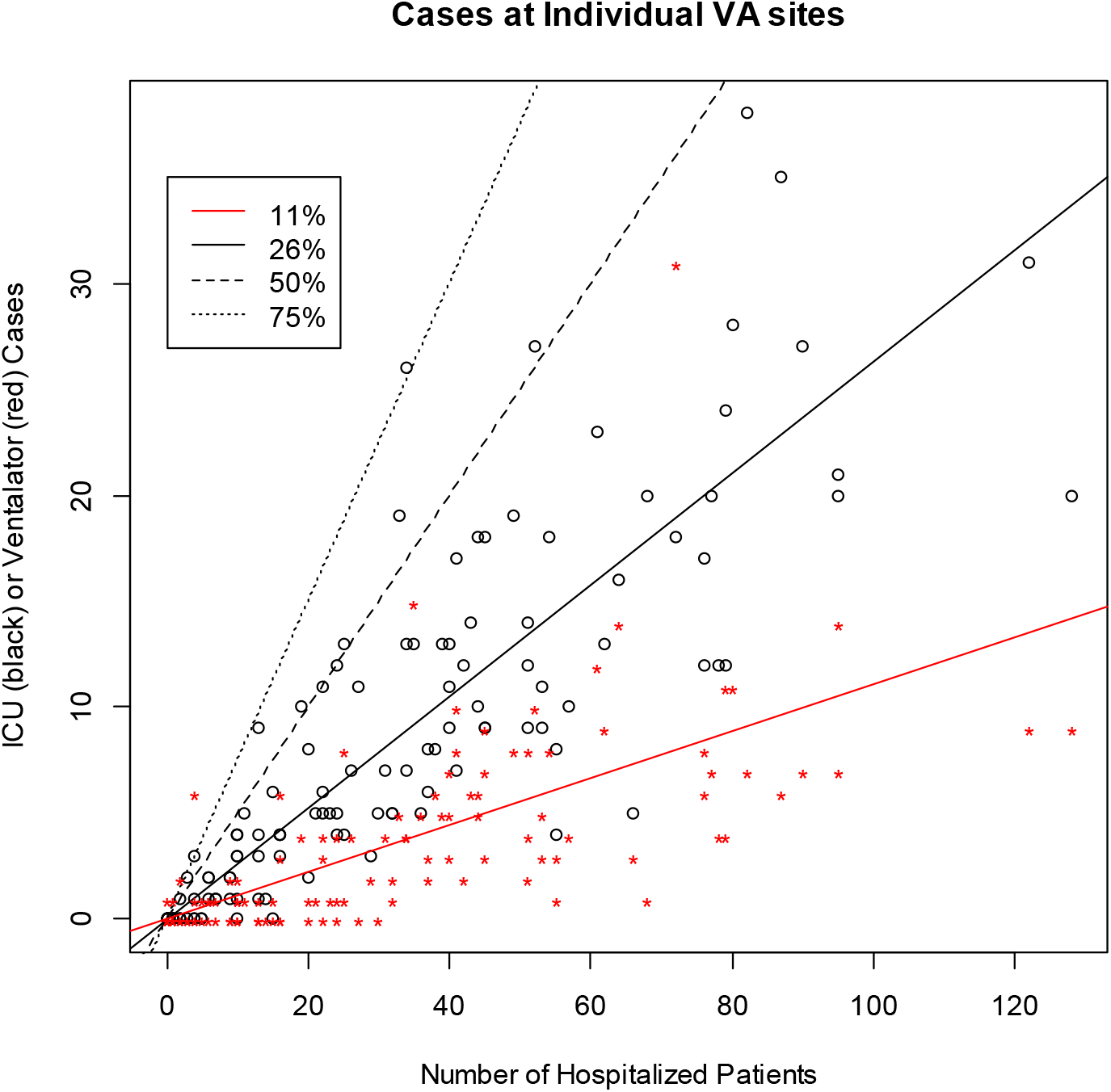
Number of cases of hospitalized, admitted to ICU, and requiring ventilator at each VA station. The solid red and black lines indicate the overall average of ventilator and ICU rates among hospitalized patients, respectively. Dashed line designates 50% rate; dotted line, 75% rate.

**Supplementary Figure 3.**
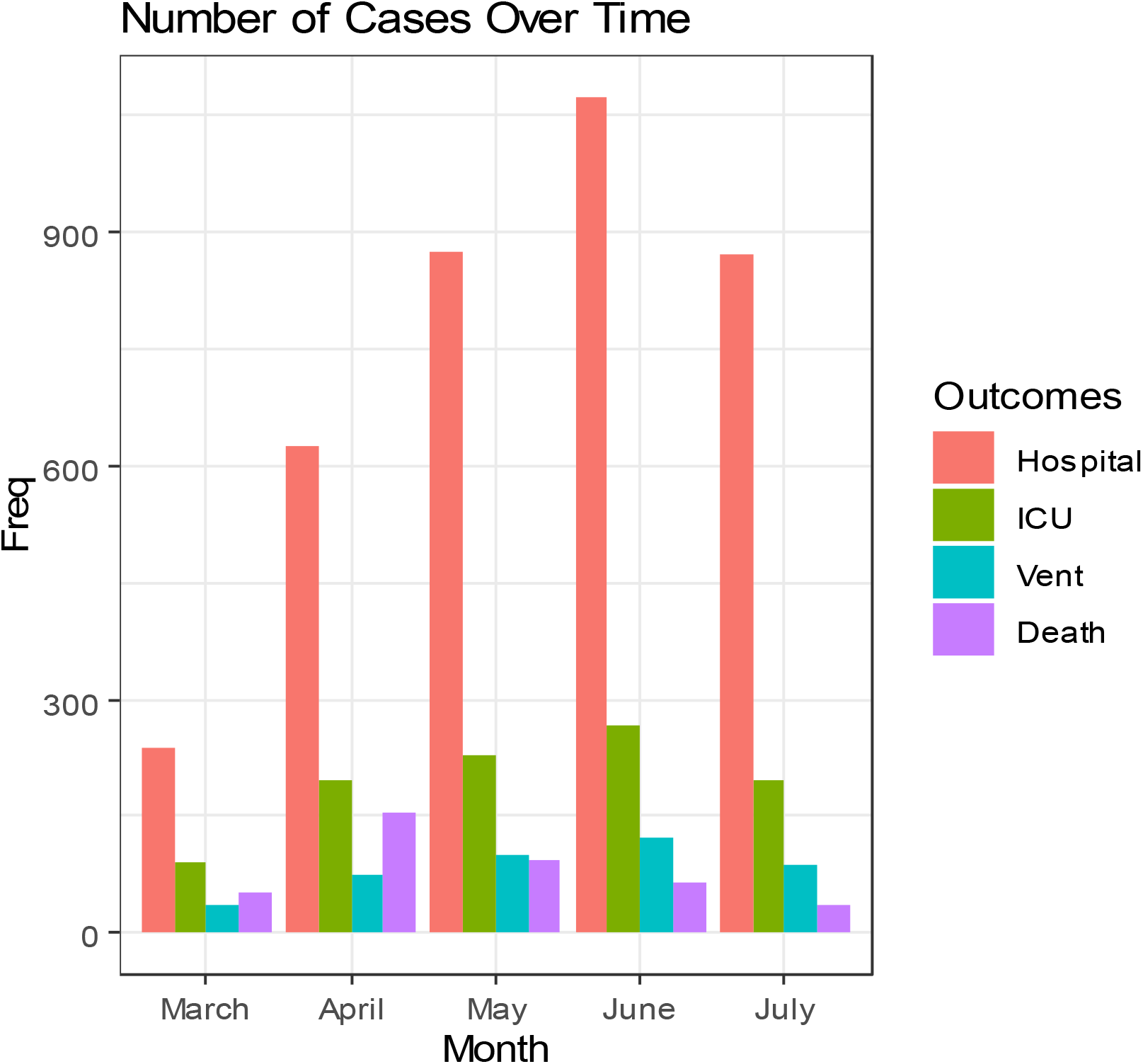
Number of cases for each outcome among patients tested for COVID-19 over time.

